# Factors associated with contraceptive failure in Uganda: Analysis of the 2016 Uganda Demographic and Health Survey

**DOI:** 10.1101/2022.02.09.22270716

**Authors:** Ruth Ketty Kisuza, Saviour Kicaber, Derrick Abila Bary, Felix Bongomin, Christopher Orach Garimoi

**Affiliations:** College of Health Sciences, Makerere University, Kampala Uganda; Department of Medical Microbiology and Immunology, Faculty of Medicine, Gulu University, Gulu, Uganda

**Keywords:** Contraceptive failure, Uganda, Demographic Health Survey (DHS), Socio-demographic Characteristics

## Abstract

**Introduction:** Sustained motivation is essential for effective use of contraceptive methods by women in low- and middle-income countries as many women are likely to abandon use of contraceptives especially when they continually experience episodes of failure. We aimed to determine contraceptive failure rates and associated factors among Ugandan women using data from the 2016 Uganda Demographic Health Survey (UDHS).

**Methods:** We analyzed data collected by the UDHS conducted in Uganda 2016. All eligible women aged 15 to 49 years at the time of the survey were enrolled. Discontinuation of contraceptive use due to failure within a 5-year period preceding the survey was the dependent variable.

**Results:** A total of 18,505 women were included in this study, 70.8% (n=5153) lived in rural areas while 56.9% (n=5153) owned a mobile phone. The mean age of the women was 29.6years (SD 7.6). The overall prevalence of contraceptive failure was 5.6%, and was higher (7.8%) among women aged 25-29 years or had completed secondary education (7.1%). The odds of contraceptive failure was 38% lower in women who had an informed choice on contraceptives compared to those who didn’t [Adjusted Odd ratio, 0.62; 95% confidence interval, 0.50 – 0.77; p< 0.001].

**Conclusion:** The burden of contraceptive failure among women of reproductive age in Uganda is substantial and significantly varied by socio-demographic characteristics.

## Introduction

Improving access to family planning (FP) services is fundamental to achieving the Sustainable Development Goals (SDGs) because it is strongly related with women’s and children’s health, poverty reduction, education, gender equality, and human rights. Access to family planning contributes up to a 44% reduction in maternal deaths [1]. Since most unplanned pregnancies and abortions occur in women who were either not using contraception or not using it consistently. Greater access to contraception and more consistent use of contraception are crucial in the reduction of unplanned pregnancies and abortions.

The reasons for discontinuation of contraceptives can be grouped by whether they represent discontinuations due to reduced need for contraception (not in need) or discontinuations while women were presumably still exposed to the risk of pregnancy and did not want to become pregnant (in need). Among discontinuations that are not in need, the most common reason given is wanting to become pregnant. Among in-need discontinuations, the most common reasons given are women that become pregnant while using the method (contraceptive failure) or those that stopped using the method because of side effects[2].

High rates of discontinuation for reasons other than the desire for pregnancy are problematic because of their association with several negative reproductive health outcomes. In countries with moderate-high contraceptive prevalence most of the unintended pregnancy is the result of contraceptive discontinuation or failure. Several studies have associated contraceptive discontinuation for reasons other than the desire to become pregnant with unmet need for contraception and induced abortion ([2–4]. Unintended pregnancy has been associated with increased risk of maternal morbidity, health behaviors during pregnancy that are associated with adverse maternal health, and adverse fetal, infant and child health outcomes [5] Additionally, unintended pregnancy has been associated with negative psychological effects on women and their children[6].

In the developing world, 74 million unintended pregnancies occur annually, and nearly a third 30%, are due to contraceptive failure among women using some type of contraceptive method (whether traditional or modern). This includes both method-related failures (i.e., failure of a method to work as expected) and user-related failures (i.e., failure due to incorrect or inconsistent use of a method)[7].

Sustained motivation is essential for effective use of contraceptive methods by women in low- and middle-income countries as many women are likely to abandon use of contraceptives especially when they continually experience episodes of failure. Assessing failure rates among demographic and socioeconomic groups is important to inform efforts to improve contraceptive information, services and use, and to minimize contraceptive failures. Additionally, detailed information on contraceptive failure rates is critical to inform improvements in provision of contraceptive information, supplies and services, which can help women and couples to use methods correctly and consistently. This study aimed to determine the contraceptive failure rates and the factors associated with contraceptive failure among Ugandan women using the 2016 Uganda Demographic Health Survey (UDHS) data.

## Methods

### Study design and setting

We analyzed data collected during the UDHS conducted in 2016. “The Demographic and Health Surveys (DHS) are internationally comparable household surveys that collect information on demographic, socioeconomic, and health-related variables among nationally representative samples of households in developing countries. Details of the DHS sampling design and strategies are described elsewhere”[8].

### Study population

In this study, we used the women recode file which included women aged 15 to 49 years at the time of the survey. We included all the women in the dataset into our analysis.

### Study variables

Discontinuation of contraceptive use due to failure within a five-year period preceding the survey was the dependent variable. The DHS does not have a variable that records discontinuation of contraceptive use due to failure. This was derived from the variable v360 that records the various reasons for discontinuation such as pregnancy, wanting to become pregnant, side effects and wanting a more effective method. In this study, a woman becoming pregnant while on contraceptives was termed as a method failure (exposure) while discontinuation due to another reason was termed as non-exposure.

The independent variables used in the study included (1) age, (2) type of place of residence (rural vs urban), (3) wealth index, (4) education attainment, (5) phone ownership, (6) internet use, (7) number of children delivered, (8) Women’s participation in decision-making for using contraceptives, and (9) work status. We also included the variable on the method of contraception that was discontinued over the 5-year period preceding the survey.

### Data management

The data used were obtained after receiving permission from the DHS program website. The dataset for the Uganda DHS 2016 was then downloaded from the DHS program website. The women’s recode file readable by STATA version 13 was selected for use.

To calculate failure rate, we created a new variable for which the denominator would be the total of the number of women who were currently using contraceptives at time of survey interview and the number of women who were not using at the time of survey because they had discontinued. The number of women who were currently using contraceptives at time of survey were obtained by converting variable v312 in the DHS into a binary variable (currently use vs. non-currently using). The number of women who were not using contraceptives at the time of the survey because they discontinued was obtained in the following way. First, we created a new variable for only women who were not currently using contraceptives at the time of the survey. This was recorded into a binary variable with 0 if a woman had not discontinued contraceptives and 1 if a woman discontinued contraceptives. The latter was used to obtain the number of women who were not using contraceptives at the time of the survey because they discontinued.

The numerator for calculating contraceptive failure rate was obtained by converting DHS variable v360 (reason for discontinuation) into a binary variable with yes (1) if reason was failure/pregnancy and no (0) if other reason for discontinuation. The numerator considered was the number of women who discontinued due failure.

### Data analysis

All the analysis in this study was performed in STATA13 and each survey dataset was analyzed separately (StataCorp LP 2013). Weighting was performed for all the descriptive statistics using the weight variable (v005) after dividing it by 1,000,000.

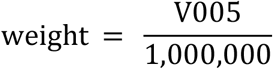

For the regression analysis in STATA13, weighting was also performed using the Primary Sampling Unit (PSU) and Strata as the variables v021 and v022 respectively in the DHS survey datasets for the surveys and weight (wgt) calculated previously. The STATA13 code below was used to apply weights before calculating means, standard deviation and 95% confidence intervals of continuous variables and regression analysis.

svyset v021 [pw = wgt], strata(v022) single unit(centered)

where pw is the probability weight (sampling weight), psu is primary sampling unit, v021 is variable for the primary sampling unit, and v022 is the variable in the DHS that indicates the strata used in the DHS surveys.

### Summary statistics

For the categorical variables, we summarized them as weighted proportions. Variables that were numeric like age were summarized as weighted means reporting standard deviations and 95% confidence intervals.

### Regression analysis

We used bivariate and multivariate logistic regression to determine the association between demographic characteristics and contraceptive failure among women. We reported crude odds ratios, adjusted odds ratios, p-values, and 95% confidence intervals. First, bivariate logistic regression was used to determine the association between contraceptive failure and the women’s demographic characteristics. Then, multivariate logistic regression was used including the variables that had a p-value of less than 0.200 i.e. (1) age group (2) number of children a woman has, and (3) Frequency of using the internet last month.

### Quality control

To ensure that we are working with the correct DHS dataset for a given year, a test analysis was performed to try to replicate the tables in the respective DHS survey reports for the various years. We created a frequency distribution table for the variable of type of place of residence for the different years in STATA13 and compared the result with what was reported in the respective DHS survey report. All results from this test analysis matched those in the DHS survey reports.

## Results

### Characteristics of the women

**Table 1** shows the socio-demographic characteristics of the sample of women aged 15-49 who had used and never used a contraceptive method within the five years before the 2016 UDHS. Among contraceptive users (n=8000), 23.4% were between age 25-29 years, only 7.6% (n=691) were adolescents (age 15-19 years) and 4.4% (n=401) were above age 45-49 years. Only 7.8% had no formal education. Majority (80.3%) of the women were working and about 70.7% lived in rural areas of Uganda. About 28.9% (n=2573) of the women belonged to the highest wealth quintile. Majority of women who used contraceptives had never been exposed to the internet in the last 12 months (89.0%), 9.6% (n= 873) had used the internet in the last 12 months. Additionally, 56.8% (n= 5153) of the women who used contraceptives owned a mobile phone. Among cultural factors, about two-thirds of the women made a joint decision with their partners on contraceptive use. 41.8% (n=3788) of the women using contraceptives made an informed choice while 58.2% (n=5273) of the women did not make an informed choice. The most common methods used and discontinued in the past 5 years were injectable (54.8%), followed by implants (10.5%).

**Table 1:**
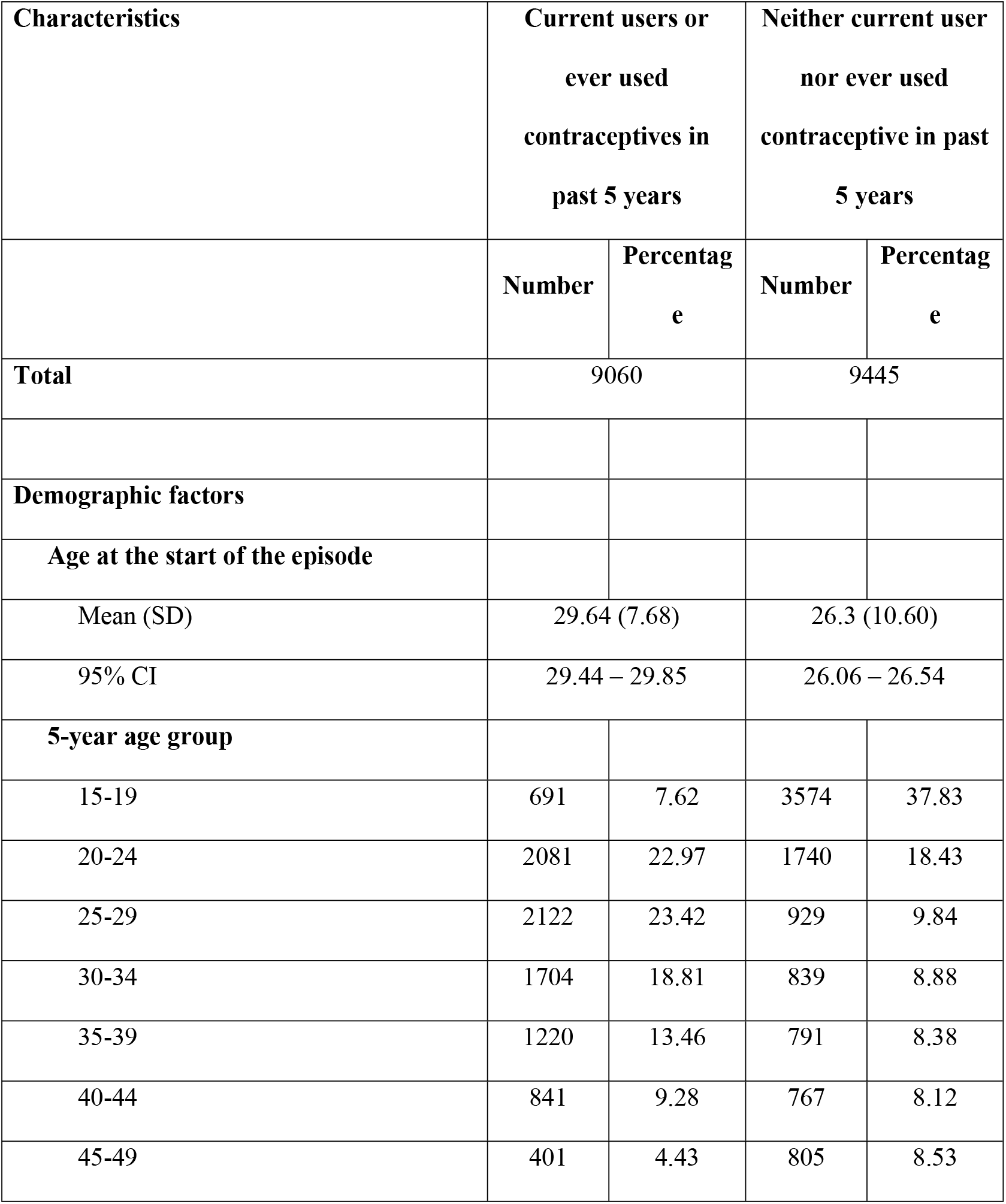
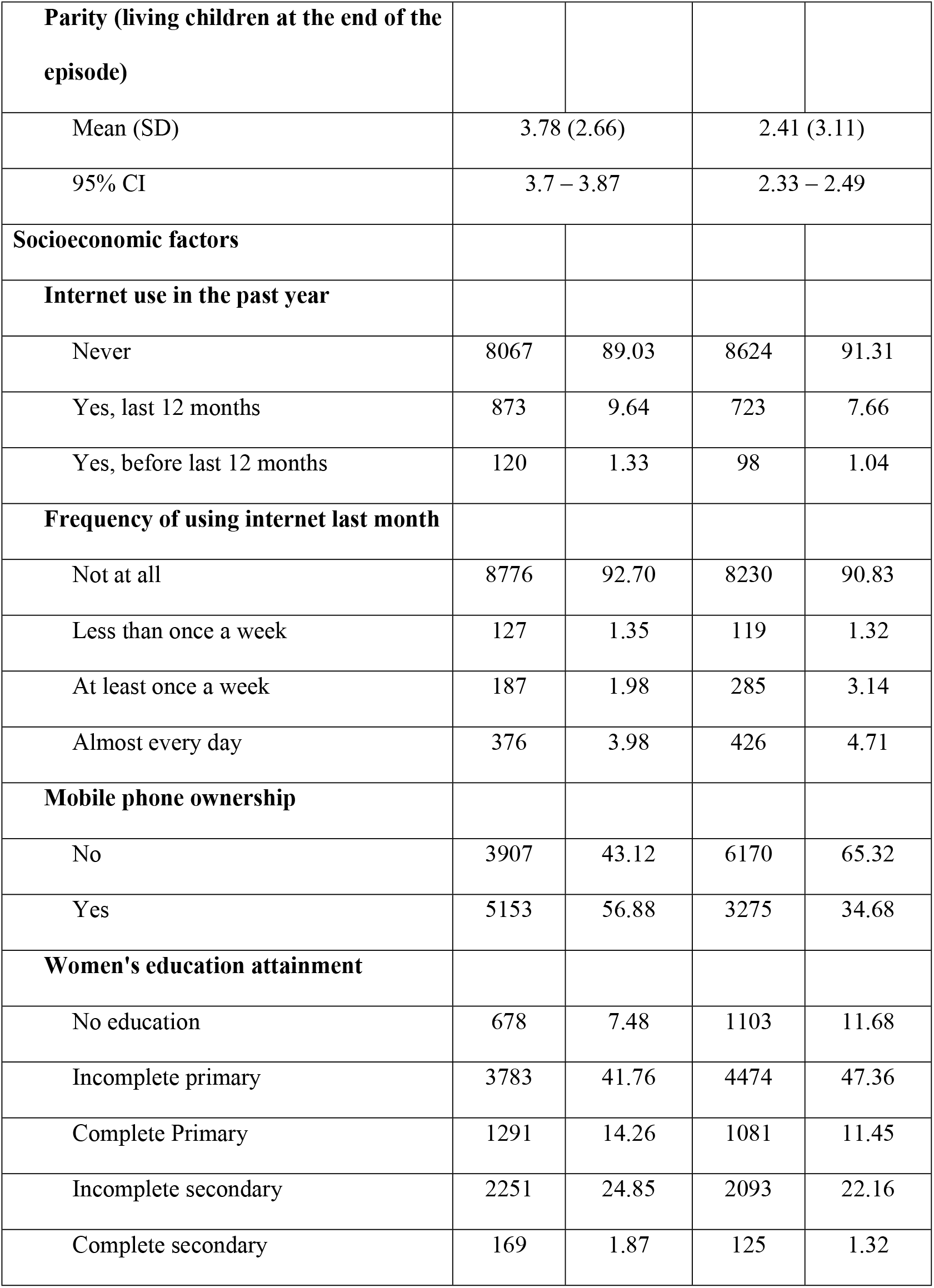
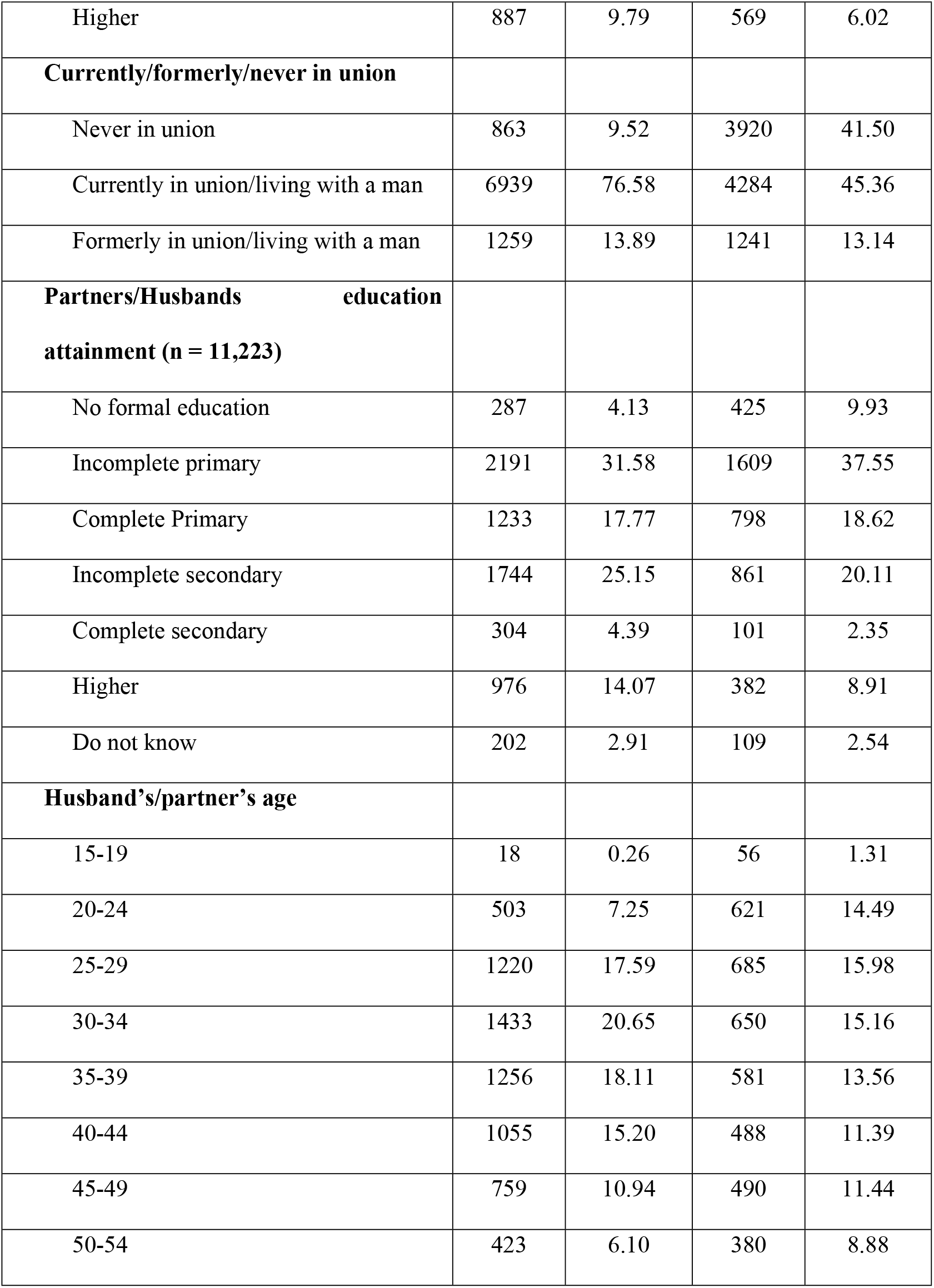
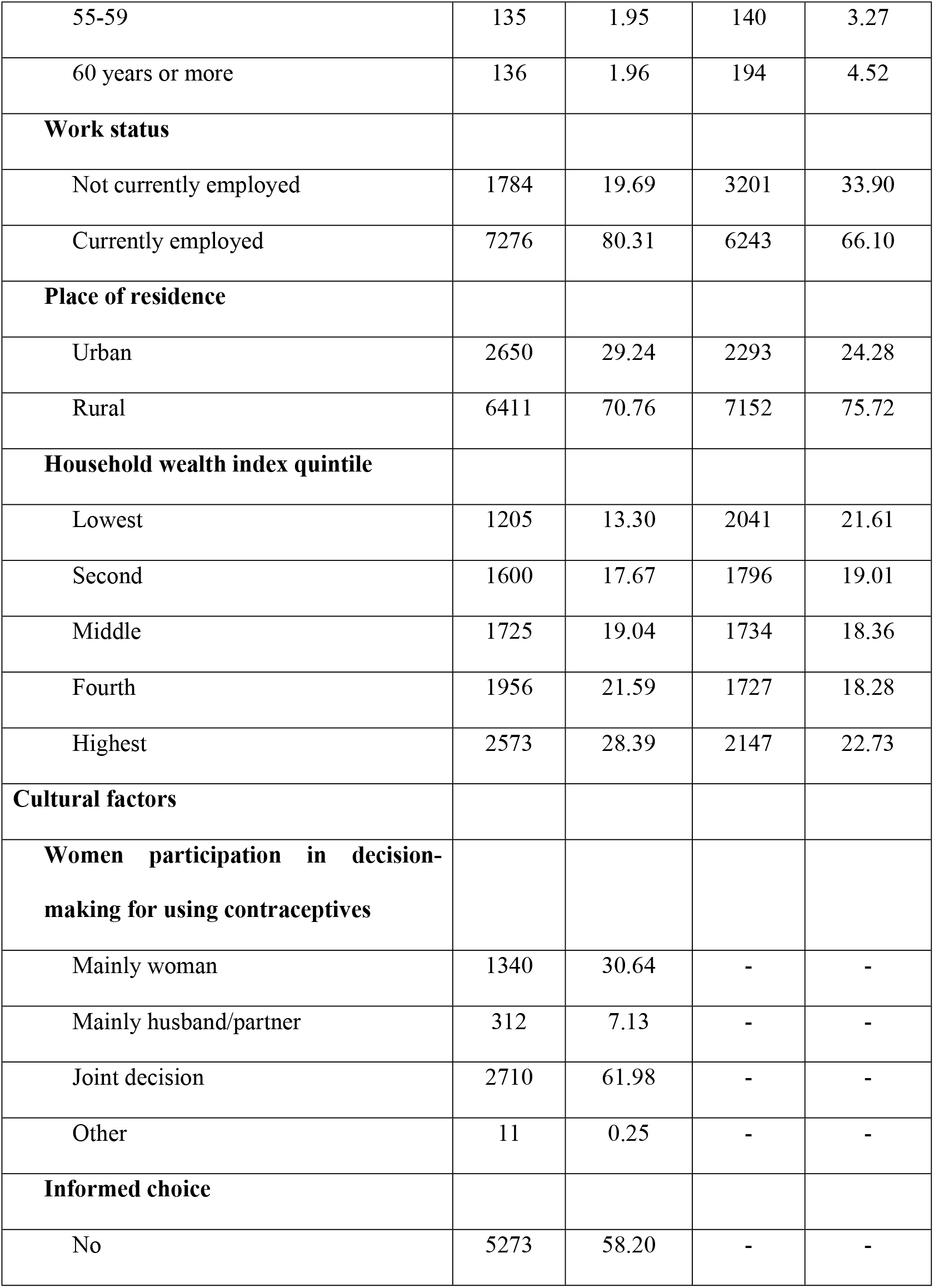
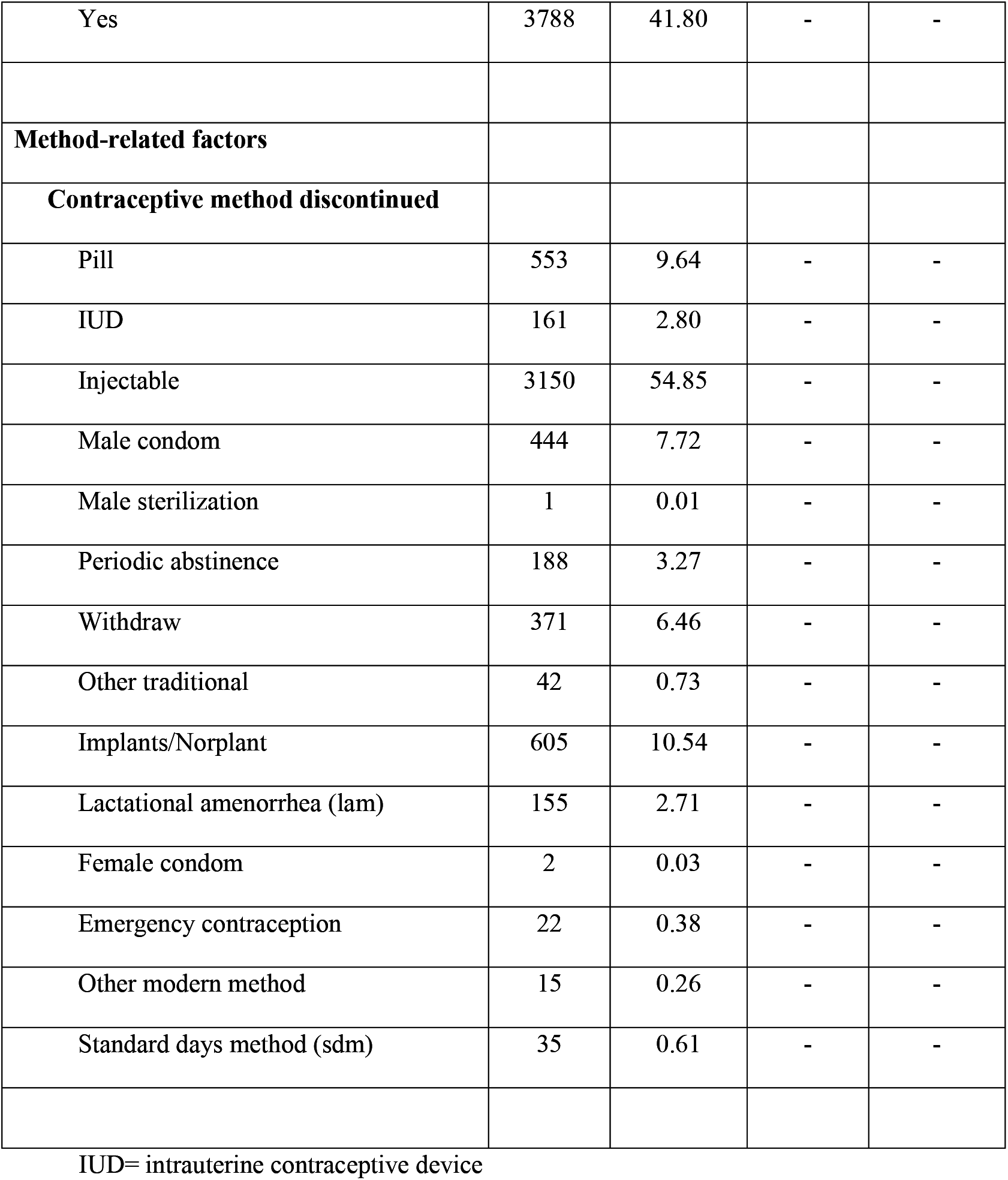
Distribution of method related, demographic, socio economic and cultural characteristics of women by status of contraceptive use.

### Prevalence of contraceptive failure

As shown in **Table 2**, the prevalence of contraceptive failure among Ugandan women aged 15-49 years during the 5-year period by demographic, socio economic and cultural factors was 5.6% (n=506). Among the demographic factors, failure rate 7.83% (n=157) was prevalent in women aged 25-29 years and having five or more children (6.35%). For socioeconomic factors, the highest prevalence rates were seen in women who used internet almost every day in the one month (7.78%, n=426), owned a mobile phone (5.72%, n=294), completed higher education (7.09%, n=63), unemployed (5.83%, n=104) and lived in a rural area (5.70%, n=364).

**Table 2:**
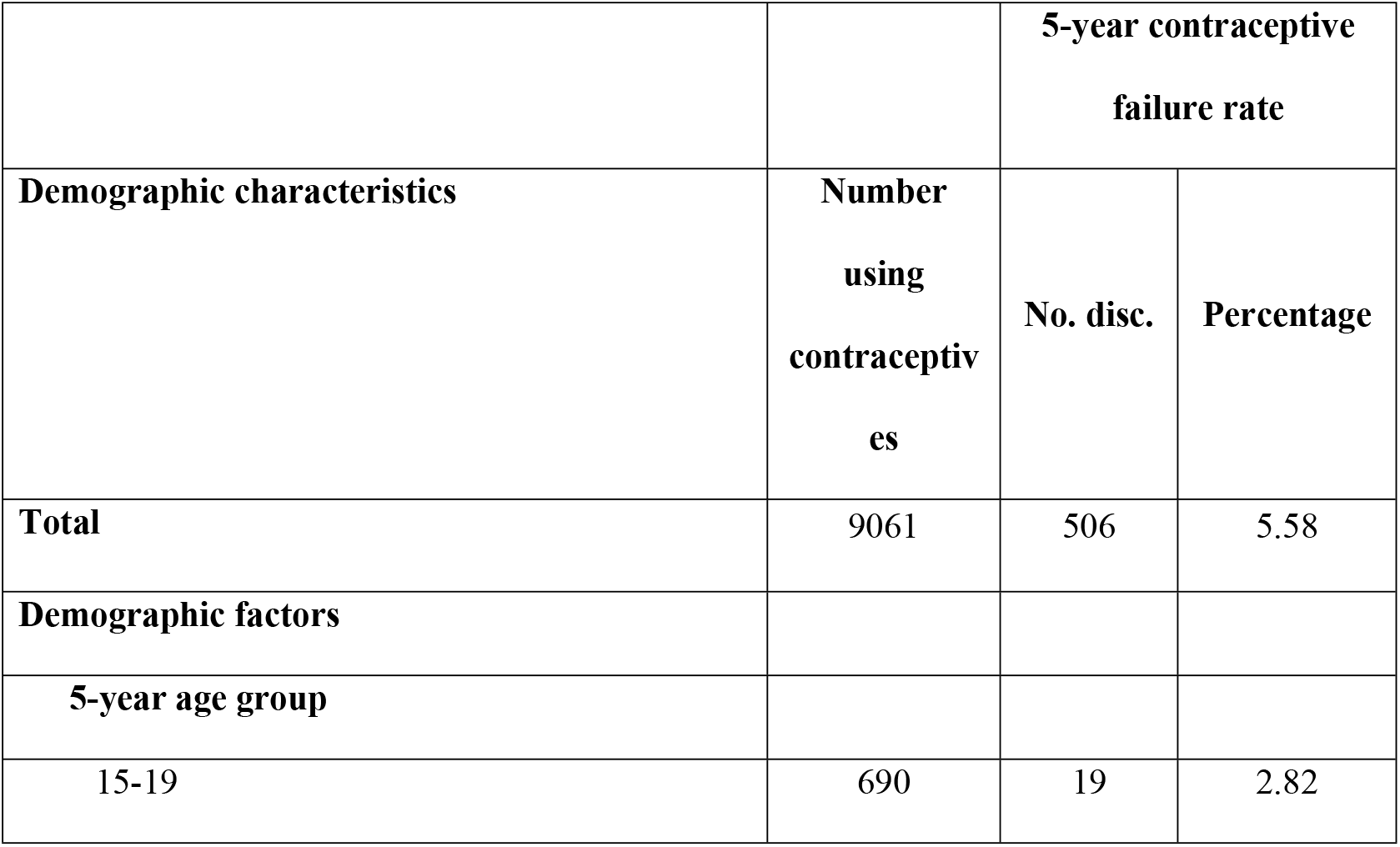
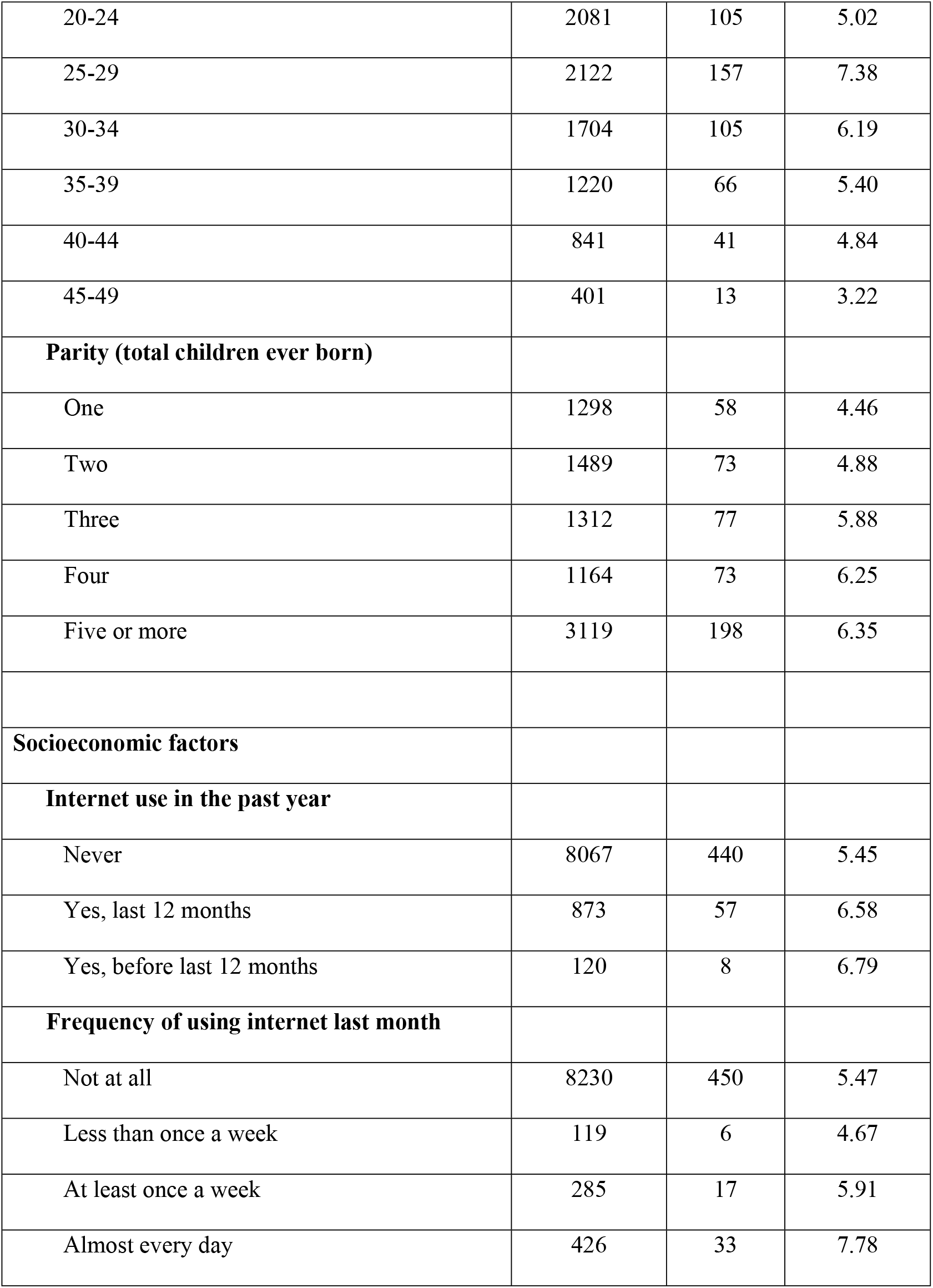
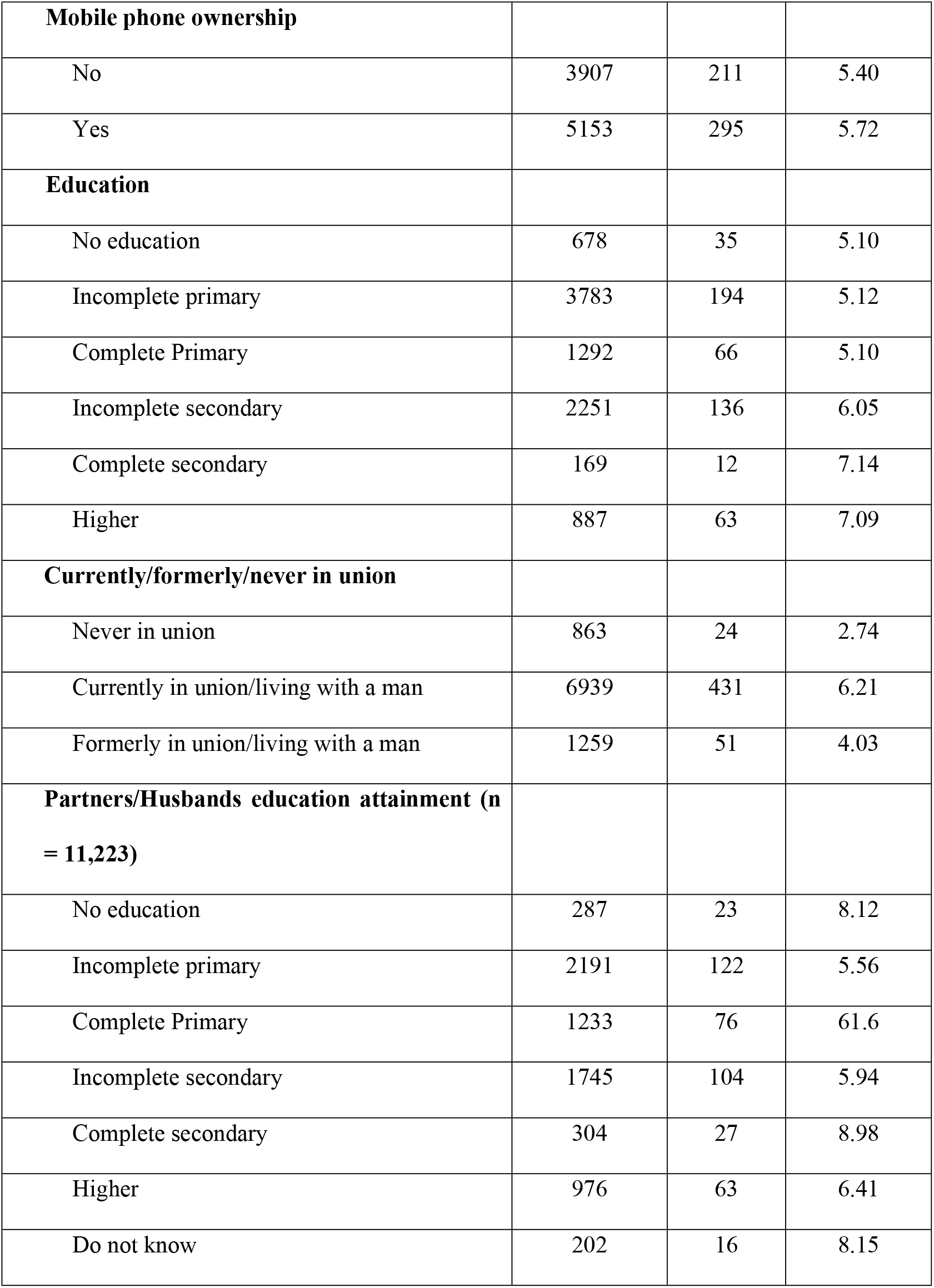
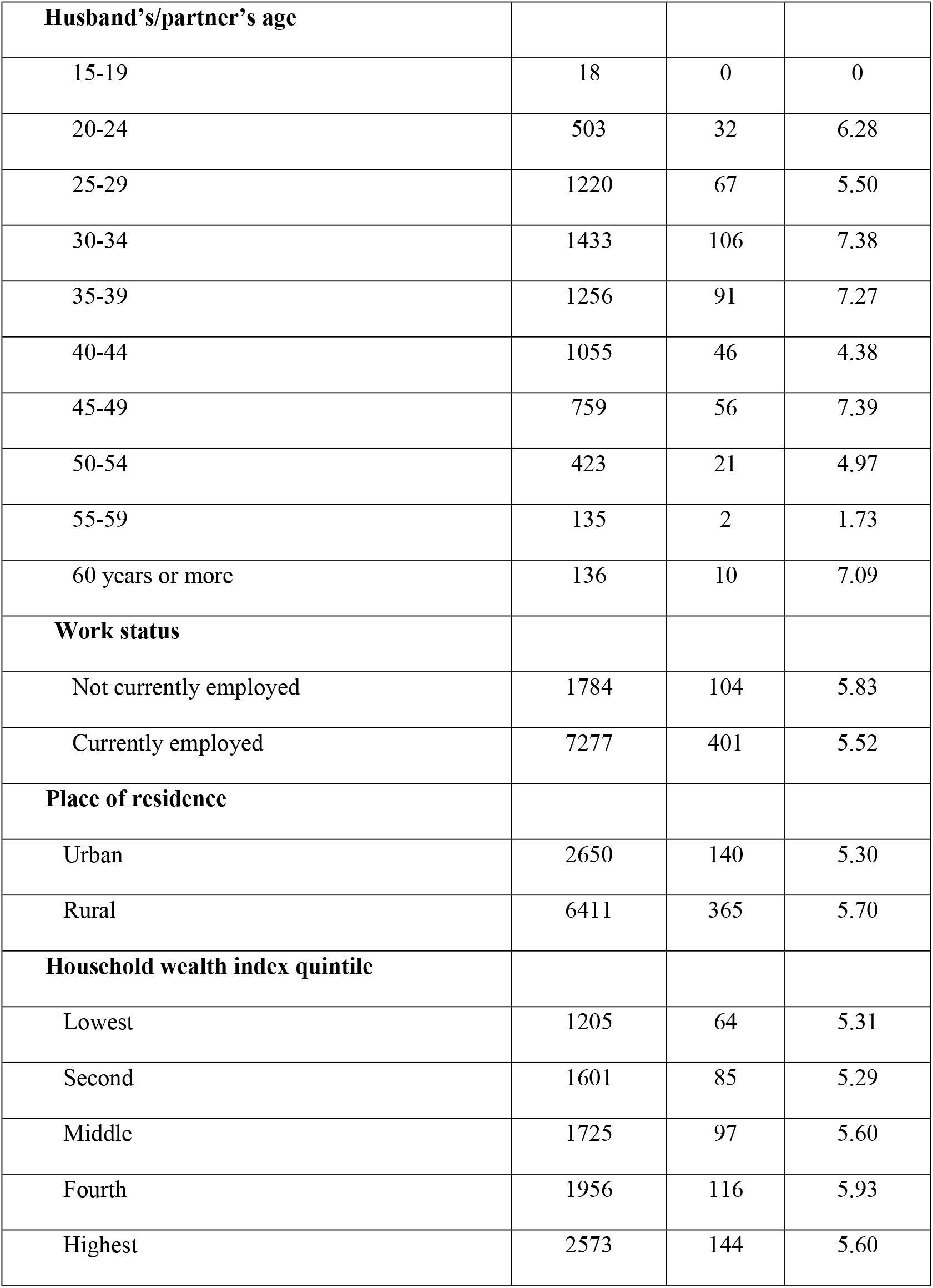
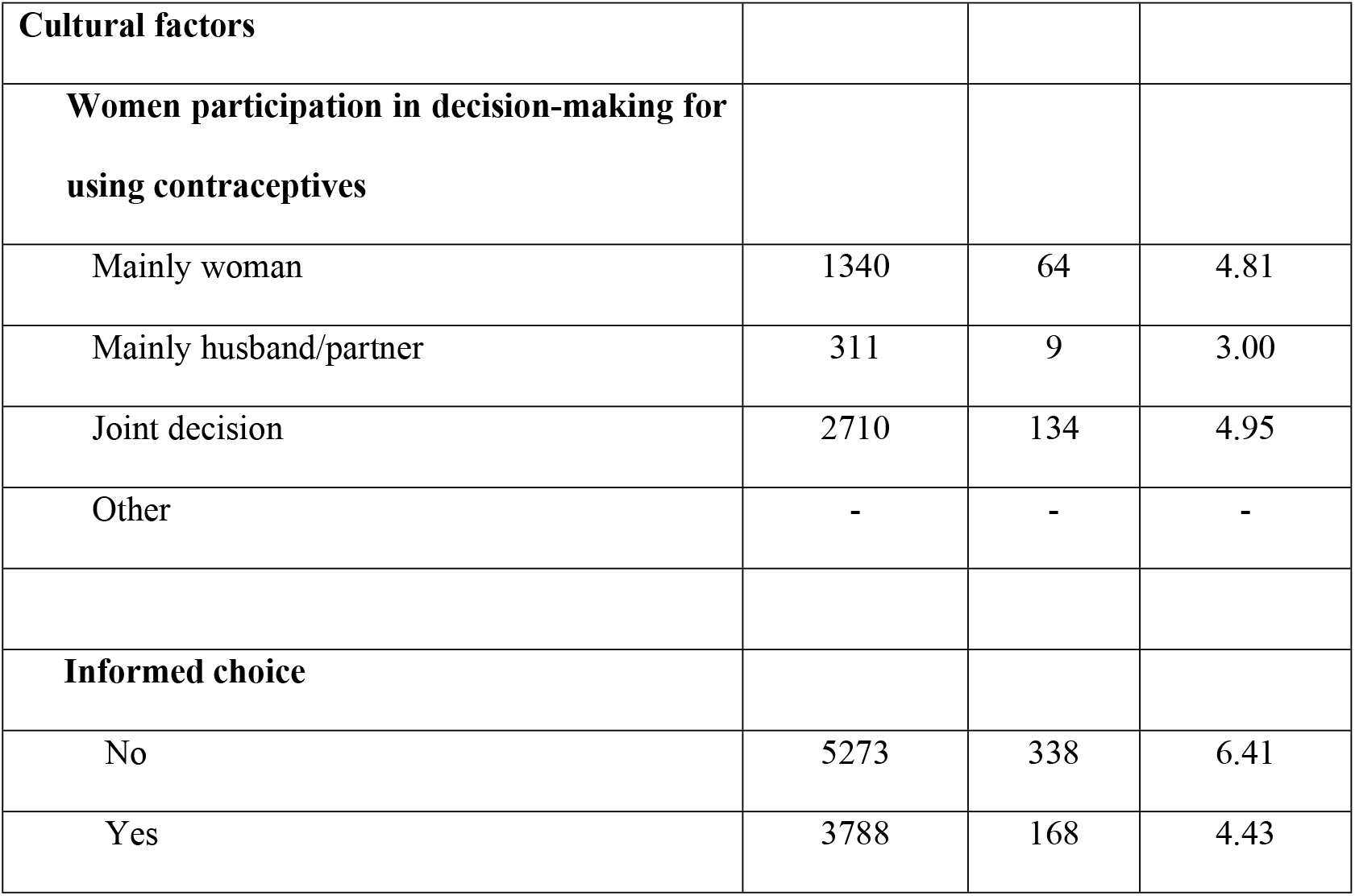
Prevalence of contraceptive failure among Ugandan women aged 15-49 years during a five-year period by demographic, socioeconomic, and cultural factors.

Among cultural factors, women who had a joint decision for contraceptive use with their partner had the highest failure of the 4.95% (n=134).Women who made an informed choice on contraceptive use had low contraceptive failure rates (4.43%, n=168) as compared to those who didn’t make an informed choice (6.41%, n=338).

**Table 3** shows the contraceptive failure rates in Uganda by method used. Generally traditional methods had the highest failure rates. Of this, withdrawal (36.30%, n=134) was the single most common reason for failure. For modern contraceptive methods, short term methods like emergency contraception (20.85%, n=5) and pills (10.68%, n=59) had higher failure rates when compared to long term modern contraceptives like IUD (1.62%, n=3) and implants (2.60%, n=15) which had very low failure rates.

**Table 3:** Contraceptive failure rates in Uganda by methods used.

|  |  |  |  |
| --- | --- | --- | --- |
| **Method-related factors** |  |  |  |
| **Contraceptive method discontinued** | **Number who used the method** | **Number failed** | **Percentage (%)** |
| Pill | 553 | 59 | 10.68 |
| IUD | 161 | 3 | 1.62 |
| Injectable | 3150 | 140 | 4.43 |
| Male condom | 444 | 31 | 6.93 |
| Male sterilization | 1 | 0 | 0 |
| Periodic abstinence | 188 | 65 | 34.87 |
| Withdraw | 371 | 134 | 36.30 |
| Other traditional | 41 | 19 | 44.78 |
| Implants/Norplant | 605 | 15 | 2.60 |
| Lactational amenorrhea<br>(lam) | 155 | 20 | 12.69 |
| Female condom | 2 | 1 | 50.00 |
| Emergency contraception | 22 | 5 | 20.85 |
| Other modern method | 15 | 2 | 13.43 |
| Standard days method (sdm) | 35 | 12 | 34.17 |

### Factors associated with contraceptive failure

From the multivariate analysis, women aged 45 to 49 years [Adjusted Odds ratio (aOR), 0.33; 95% confidence interval (95CI), 0.12 – 0.94; p value = 0.038] were less likely to experience a contraceptive failure compared to those aged 15 to 19 years. Women who had an informed choice on contraceptives were less likely to experience a failure [aOR, 0.62; 95CI, 0.50 – 0.77; p-value < 0.001] compared to those who did not have informed choice (**Table 4**).

**Table 4:** Factors associated with contraceptive failure among women in Uganda.

|  | Bivariate analysis |  | Multivariate analysis |  |
| --- | --- | --- | --- | --- |
| Demographic characteristics | Crude Odds Ratio<br>(95% CI) | P - value | Adjusted Odds<br>Ratio<br>(95% CI) | P - value |
| Demographic factors |  |  |  |  |
| 5-year age group |  |  |  |  |
| 15-19 | 1 |  | 1 |  |
| 20-24 | 1.73 (1.04 – 2.86) | 0.034 | 1.28 (0.66 – 2.49) | 0.464 |
| 25-29 | 2.61 (1.59 – 4.27) | < 0.001 | 1.56 (0.75 – 3.25) | 0.234 |
| 30-34 | 2.19 (1.32 – 3.63) | 0.002 | 0.98 (0.45 – 2.16) | 0.967 |
| 35-39 | 2.05 (1.22 – 3.46) | 0.007 | 0.83 (0.36 – 1.90) | 0.664 |
| 40-44 | 1.56 (0.88 – 2.76) | 0.127 | 0.53 (0.22 – 1.3) | 0.166 |
| 45-49 | 1.05 (0.496 – 2.211) | 0.903 | **0.33 (0.12 – 0.94)** | **0.038** |
| **Parity (total children ever born)** |  |  |  |  |
| No child | 1 |  | 1 |  |
| One child | 1.58 (0.93 – 2.66) | 0.089 | 0.68 (0.34 – 1.36) | 0.277 |
| Two children | 1.45 (0.87 – 2.44) | 0.157 | 0.66 (0.33 – 1.32) | 0.239 |
| Three children | 1.80 (1.08 – 3.01) | 0.024 | 0.79 (0.39 – 1.62) | 0.522 |
| Four children | 1.895 (1.131 – 3.175) | 0.015 | 0.92 (0.43 – 1.93) | 0.818 |
| Five or more children | 2.16 (1.35 – 3.47) | 0.001 | 1.70 (0.80 – 3.61) | 0.166 |
| **Socioeconomic factors** |  |  |  |  |
| **Internet use in the past year** |  |  |  |  |
| Never | 1 |  | - |  |
| Yes, last 12 months | 1.08 (0.78 – 1.49) | 0.637 | - | - |
| Yes, before last 12 months | 1.42 (0.70 – 2.88) | 0.329 | - | - |
| **Frequency of using internet last month** |  |  |  |  |
| Not at all | 1 |  | - |  |
| Less than once a week | 0.73 (0.28 – 1.88) | 0.512 | - | - |
| At least once a week | 1.46 (0.89 – 2.398) | 0.136 | - | - |
| Almost every day | 1.01 (0.64 – 1.59) | 0.971 | - | - |
| **Mobile phone ownership** |  |  |  |  |
| No | 1 |  | - |  |
| Yes | 0.99 (0.82 – 1.19) | 0.918 | - | - |
| **Education** |  |  |  |  |
| No education | 1 |  | - |  |
| Incomplete primary | 1.01 (0.698 – 1.454) | 0.969 | - | - |
| Complete Primary | 1.09 (0.72 – 1.65) | 0.678 | - | - |
| Incomplete secondary | 1.18 (0.799 – 1.73) | 0.410 | - | - |
| Complete secondary | 1.32 (0.63 – 2.77) | 0.461 | - | - |
| Higher | 1.20 (0.77 – 1.89) | 0.416 | - | - |
| **Currently/formerly/never in union** |  |  |  |  |
| Never in union | 1 |  |  |  |
| Currently in union/living with a man | 2.15 (1.41 – 3.28) | < 0.001 | - |  |
| Formerly in union/living with a man | 1.39 (0.84 – 2.30) | 0.205 | - |  |
| **Partners/Husbands education attainment (n = 11,223)** |  |  |  |  |
| No education | 1 |  |  |  |
| Incomplete primary | 0.66 (0.42 – 1.04) | 0.074 | - | - |
| Complete Primary | 0.76 (0.47 – 1.22) | 0.253 | - | - |
| Incomplete secondary | 0.68 (0.42 – 1.09) | 0.110 | - | - |
| Complete secondary | 1.06 (0.58 – 1.94) | 0.843 | - | - |
| Higher | 0.76 (0.46 – 1.25) | 0.278 | - | - |
| Do not know | 0.90 (0.45 – 1.81) | 0.772 | - | - |
| **Husband's/partner's age** |  |  |  |  |
| 15-19 | 1 |  | 1 |  |
| 20-24 | 2.93 (0.17 – 49.43) | 0.456 | 2.75 (0.16 – 47.27) | 0.486 |
| 25-29 | 2.69 (0.16 – 44.71) | 0.491 | 1.99 (0.11 – 34.47) | 0.638 |
| 30-34 | 3.81 (0.23 – 63.21) | 0.350 | 2.43 (0.14 – 42.32) | 0.543 |
| 35-39 | 3.39 (0.20 – 56.34) | 0.394 | 2.26 (0.13 – 39.68) | 0.577 |
| 40-44 | 2.49 (0.15 – 41.64) | 0.524 | 1.75 (0.098 – 31.00) | 0.704 |
| 45-49 | 3.42 (0.21 – 57.14) | 0.392 | 2.78 (0.16 – 49.49) | 0.487 |
| 50-54 | 2.66 (0.16 – 45.18) | 0.499 | 2.5 (0.14 – 45.56) | 0.537 |
| 55-59 | 1.4 (0.07 – 27.98) | 0.826 | 1.46 (0.07 – 31.20) | 0.810 |
| 60 years or more | 3.34 (0.19 – 59.89) | 0.412 | 3.02 (0.16 – 57.77) | 0.463 |
| **Work status** |  |  |  |  |
| Not currently employed | 1 |  | - |  |
| Currently employed | 1.02 (0.80 – 1.31) | 0.849 | - | - |
| **Place of residence** |  |  |  |  |
| Urban | 1 |  | 1 |  |
| Rural | 1.03 (0.83 – 1.27) | 0.803 | 1.04 (0.8 – 1.4) | 0.789 |
| **Household wealth index quintile** |  |  |  |  |
| Lowest | 1 |  | - |  |
| Second | 1.06 (0.78 – 1.45) | 0.712 | - | - |
| Middle | 0.98 (0.71 – 1.34) | 0.901 | - | - |
| Fourth | 1.05 (0.77 – 1.43) | 0.747 | - | - |
| Highest | 0.95 (0.70 – 1.29) | 0.760 | - | - |
| **Cultural factors** |  |  |  |  |
| **Women participation in decision-making for using contraceptives** |  |  |  |  |
| Mainly woman. | 1 |  | - |  |
| Mainly husband/partner | 0.75 (0.38 – 1.46) | 0.399 | - | - |
| Joint decision | 1.01 (0.73 – 1.399) | 0.933 | - | - |
| Other | 1.27 (0.07 – 22.20) | 0.872 | - | - |
| **Informed choice** |  |  |  |  |
| No | 1 |  | 1 |  |
| Yes | 0.65 (0.53 – 0.79) | < 0.001 | 0.62 (0.50 – 0.77) | < **0.001** |

## Discussion

In this study, we aimed to establish and describe the prevalence of contraceptive failure amongst women of reproductive age in Uganda. We used data from the demographic and health survey conducted in Uganda in 2016. From this study, it is evident that the prevalence of contraceptive failure varies by age, type of place of residence, phone ownership, education attainment and wealth index. The highest failure rates were seen in women who use periodic abstinence and withdraw as methods of contraception. Finally, women who had delivered four or more children were more likely to experience contraceptive failure when compared with those who had not delivered any child.

The study shows that women with access to the internet were more likely to experience contraceptive failure compared to women with no access to the internet. This could be because the women with access to the internet can easily access information on the different contraceptive methods and are therefore more likely to stop or switch to better contraceptive methods when they experience side effects [9]. Additionally, women who were provided with adequate information about a particular contraceptive method and given an opportunity to make an informed choice were less likely to experience contraceptive failure than women who did not make an informed choice. This could be because a woman who made an informed choice had adequate information about the contraceptive method of her choice and was able to effectively deal with its side effects without necessarily halting contraceptive use [10,11]. This finding suggests that more time should be dedicated by clinicians to providing information on contraceptives to women before use.

In this study, failure rates by contraceptive methods were highest in women who used traditional methods than in those who used modern contraceptive methods. For modern contraceptive methods, women using short term methods like pills and Injectables were more likely to experience failure than those using long term contraceptive methods. This could be because of inconsistency when using short term contraceptive methods which exposes women to increased failure rates. Similar findings were reported in [2,12,13]. Thus, information and counselling should be emphasized for short term method users to prevent failure.

At multivariate analysis, the most important factor associated with contraceptive failure was a woman’s age. Women above 45 years of age had lower contraceptive failure rates than women below 40 years. This could be because women above 40 years are more likely than younger women to desire a permanent form of contraception[14,15]. Additionally, older women of reproductive age have lower rates of contraceptive failure than younger women and this could be explained by lower fecundity, less frequent sexual intercourse and higher compliance with contraceptive regimens as evidenced from previous studies [16].

This study used retrospective data from the DHS. The DHS follows a rigorous methodology employing a two-stage cluster randomized sampling. We used sample weights to provide estimates reported in this study. However, we acknowledge that contraceptive failure could best be studied using cohort studies. But this study provides further information about the current state of contraceptive failure among women of reproductive age in Uganda.

## Conclusion

Our study found that there was a high burden of contraceptive failure among women of reproductive age in Uganda that and varies by socio-demographic characteristics. Traditional contraceptive methods including periodic abstinence and withdrawal are associated with higher failure rates.

## Data Availability

All data sets used in this manuscript are available in the DHS program website.

## Declarations

### Ethics approval and consent to participate

Ethical approval for the Demographic and Health Surveys is obtained from relevant research ethical approval committees and research regulatory bodies before data collection. We obtained permission to use the DHS survey datasets from the DHS program website. No personally identifiable information is available in the used datasets.

### Consent for publication

Not Applicable

### Availability of data and materials

The datasets generated and/or analyzed during the current study are available in the Demographic and Health Survey program website, https://dhsprogram.com/data/available-datasets.cfm and can be accessed after obtaining approval.

### Competing interests

The authors declare that they do not have any competing interests.

### Funding

The authors did not receive any funding for this work.

### Authors’ contributions

DBA and RKK conceived the study. DBA and RKK requested data from the DHS program website, performed statistical analysis, interpretation of data and spearheaded the writing of the manuscript. RKK and DBA discussed the relevant results. DBA, RKK, SK, FB and CGO contributed to the writing of the manuscript, read, and approved the final manuscript.

## Acknowledgements

We thank the Demographic and Health Survey (DHS) program for granting us access to the DHS datasets used in this study

## Notes

### Competing Interest Statement

The authors have declared no competing interest.

### Funding Statement

The author(s) received no specific funding for this work

### Author Declarations

Ethical approval for the studies that generated this data was obtained from relevant research ethics bodies in Uganda.

## References

1. Starbird E, Norton M, Marcus R. Investing in family planning: Key to achieving the sustainable development goals. Glob Heal Sci Pract. 2016;4: 191–210. doi:10.9745/GHSP-D-15-00374

2. Ali MM, Cleland JG, Shah IH, Organization WH. Causes and consequences of contraceptive discontinuation: evidence from 60 demographic and health surveys. 2012.

3. Black KI, Gupta S, Rassi A, Kubba A. Why do women experience untimed pregnancies? A review of contraceptive failure rates. Best Pract Res Clin Obstet Gynaecol. 2010;24: 443–455.

4. Polis CB, Bradley SEK, Bankole A, Onda T, Croft T, Singh S. Contraceptive Failure Rates in the Developing World: An Analysis of Demographic and Health Survey Data in 43 Countries. Contraception. 2016;94: 11–17. Available: https://linkinghub.elsevier.com/retrieve/pii/S0010782416001037

5. Yazdkhasti M, Pourreza A, Pirak A, Abdi F. Unintended pregnancy and its adverse social and economic consequences on health system: a narrative review article. Iran J Public Health. 2015;44: 12.

6. Gipson JD, Koenig MA, Hindin MJ. The effects of unintended pregnancy on infant, child, and parental health: a review of the literature. Stud Fam Plann. 2008;39: 18–38.

7. Polis C, Bradley SEK, Bankole A, Onda T, Croft TN, Singh S. Contraceptive failure rates in the developing world: an analysis of demographic and health survey data in 43 countries. 2016.

8. Corsi DJ, Neuman M, Finlay JE, Subramanian S V. Demographic and health surveys: a profile. Int J Epidemiol. 2012;41: 1602–1613.

9. Samosir OB, Kiting AS, Aninditya F. Role of information and communication technology and women’s empowerment in contraceptive discontinuation in Indonesia. J Prev Med Public Heal. 2020;53: 117.

10. Pradhan MR, Patel SK, Saraf AA. Informed choice in modern contraceptive method use: pattern and predictors among young women in India. J Biosoc Sci. 2020;52: 846–859.

11. Hilger DJ, Raviele KM, Hilgers TA. Hormonal contraception and the informed consent. Linacre Q. 2018;85: 375–384.

12. Bradley SEK, Schwandt H, Khan S. Levels, trends, and reasons for contraceptive discontinuation. DHS Anal Stud. 2009;20.

13. Westhoff CL, Heartwell S, Edwards S, Zieman M, Stuart G, Cwiak C, et al. Oral contraceptive discontinuation: do side effects matter? Am J Obstet Gynecol. 2007;196: 412–e1.

14. Alemayehu M, Belachew T, Tilahun T. Factors associated with utilization of long acting and permanent contraceptive methods among married women of reproductive age in Mekelle town, Tigray region, north Ethiopia. BMC Pregnancy Childbirth. 2012;12: 1–9.

15. Azmoude E, Behnam H, Barati-Far S, Aradmehr M. Factors affecting the use of long-acting and permanent contraceptive methods among married women of reproductive age in East of Iran. Women’s Heal Bull. 2017;4: 1–8.

16. Allen RH, Cwiak CA, Kaunitz AM. Contraception in women over 40 years of age. CMAJ. 2013;185: 565–573.

